# Improving emergency department patient-doctor conversation through an artificial intelligence symptom taking tool: an action-oriented design pilot study

**DOI:** 10.1101/2020.11.13.20230953

**Authors:** Justus Scheder-Bieschin, Bibiana Blümke, Erwin de Buijzer, Fabian Echterdiek, Júlia Nacsa, Marta Ondresik, Matthias Ott, Gregor Paul, Tobias Schilling, Anne Schmitt, Paul Wicks, Stephen Gilbert

## Abstract

**IMPORTANCE:** Communication between patients and healthcare professionals is frequently challenging in the crowded emergency department (ED), with few opportunities to develop rapport or empathy. Digital tools for patients and physicians have been proposed as helpful but their utility is not established.

**OBJECTIVE:** To evaluate a patient-facing digital symptom and history taking, as well as handover tool in the waiting room.

**DESIGN:** A two-phase, questionnaire-based quality improvement study. Phase I observations guided iterative improvement, which was then further evaluated in Phase II.

**SETTING:** ED of a German tertiary referral and major trauma hospital providing interdisciplinary treatment for an average of 120 patients daily.

**PARTICIPANTS:** All patients who were willing/able to provide consent, excluding patients: (i) with severe injury/illness requiring immediate treatment; (ii) with traumatic injury; (iii) incapable of completing a health assessment; or, (iv) under 18 years old. Of 1699 patients presenting to the ED, 815 were eligible based on triage level. With available recruitment staff, 135 were approached, of whom 81 were included in the study.

**INTERVENTION/OBSERVATION:** Patients entered information into the tool, which generated a handover report to be accessed via a clinician dashboard. All users completed evaluation questionnaires. Clinicians were trained to observationally assess the tool as a prototype, without relying upon it for clinical care.

**MAIN OUTCOMES AND MEASURES:** Patient and clinician Likert scale ratings of tool performance.

**RESULTS:** Respondents were strongly positive in endorsing the tool’s usefulness in facilitating conversation (75% of patients, 73% physicians, 100% nurses). Nurses judged the tool as potentially time saving, whilst physicians assessed it as time saving only in some ED medical specialisms (e.g. Surgery). Patients understood the tool questions and reported high usability. The proportion of patients, physicians and nurses who would recommend the tool was 78%, 53% and 76%.

**CONCLUSIONS AND RELEVANCE:** The system has clear potential to improve patient-HCP interaction and make efficiency savings in the ED. Future research and development will extend the range of patients for which the history collection has clinical utility.

**Key Points:** *Question:* Can a patient-facing digital symptom and clinical history taking tool provide conversational support, aid in symptom taking, facilitate record keeping, and lead to improved rapport between patients, physicians and nurses in the emergency department (ED)?

*Findings:* Acceptability was high, with improved rapport experienced 90% of the time for patients, 73% for physicians and 100% for nurses. Nurses assessed the tool as having workflow benefit through potential time saving. Physicians assessed the current tool design as providing time saving in certain ED medical specialisms including Surgery.

*Meaning:* The patient-facing tool for symptom and history taking provided meaningful conversation support and showed potential for efficiency savings, however, further research and testing is required before time savings can be consistently delivered to ED clinicians across the range of relevant ED medical specialisms.

## Introduction

The emergency department is, by definition, a high-stress environment, where it is critical that the health care provider’s (HCP’s) time is used optimally. As a result, effective communication and establishing empathy with patients (and colleagues) can be challenging.^1^ There is increasing recognition that hospital emergency departments (EDs) face numerous challenges related to crowding, a problem likely to continue for the foreseeable future.^2^ Barriers to effective communication most frequently include contextual factors, such as time pressures caused by a full waiting room and urgent cases above capacity.^1^ More subtle and pervasive systemic factors include process limitations and interpersonal parameters, such as societal and health disparities.^3^ It has been proposed that appropriately designed artificial intelligence (AI) based systems could reduce ED documentation errors (improving patient safety ^4–8^) and free up HCP time, which could potentially be used to improve efficiency and have more time to build rapport with patients.^8^

One such digital symptom assessment app is Ada (Ada Health GmbH, Berlin, Germany), which utilizes a probabilistic reasoning engine with an adaptive question flow to collect demographic information, medical history and symptoms. A previous study found that patients using Ada’s tool in a primary care waiting room found it helpful and easy to use. ^9^ If appropriately adapted to the setting, the benefits of a digital ED history taking tool could assist nurse-led triage in the waiting room and/or assessment and treatment by ED physicians.

### The aim of this study

In this study we evaluated a prototype digital history/symptom taking and handover system, which included a patient-facing tool for symptom and history taking. Through the patient’s interaction with this, an HCP clinical handover report was generated and displayed to the physician and/or nurse, via an HCP-facing tool. Our hypothesis was that some of the rapport, communication, documentation and time pressures in the crowded ED could be alleviated by the use by patients of a modified digital symptom assessment tool in the waiting room with transfer (handover) of a symptom assessment report to the HCPs in the ED. This pilot study was based on the approach of Furaijat et al.^10^, in which the quality improvement delivered by a prototype patient facing system was assessed in two study stages. Phase I, the action-oriented phase, included the initial implementation of the patient- and HCP-tools (version V1), and their evaluation by all users. The feedback on performance from Phase I was used to create a modified system (version V2), which was then further evaluated in Phase II. Patients, physicians and nurses quantitatively evaluated the two system versions in terms of their usability and usefulness in facilitating patient-HCP conversation and rapport formation in the ED setting. For HCP-users, we also explored the helpfulness of the medical information provided at handover and the potential of the system to save HCP time.

## Methods

### Design: overall study approach and study type

Phase I followed an action-oriented approach ^10^ which involved close collaboration between researchers and HCPs. Modifications suggested by users that could be implemented within the project timeframe were then used to develop a V2 prototype, and at a switch-over point, the V1 tool was replaced by the V2 tool, which was then fixed and remained unchanged during Phase II. There were no study design changes made between Phase I and II.

This study explored the potential for ED service quality improvement through the introduction of an augmentative prototype history and symptom taking tool. Questionnaire-based user perceptions on the potential for the system were collected alongside qualitative observations. No interventional patient outcomes measures were recorded. The system was not closely integrated into clinical workflows during the study, and it did not replace standard practice in the ED. HCPs were trained not to rely on the prototype system for formal or definitive symptom-taking support during the study. There was nonetheless a theoretical risk to patients from the study of either inappropriate prolongation of the waiting time or through inappropriate reliance of HCPs on the system. For this reason, a designated physician (JSB) made a safety analysis for each patient by examining the handover report and the patient’s clinical history in the electronic health record and adjudicated if there was any evidence of inappropriate physician guidance. This study was conducted in accordance with requirements of the SQUIRE ^11^, SPIRIT-AI and eCONSORT ^12^ guidelines, as described in the checklist (**Supplementary Table 1**).

### Description of the prototype digital symptom/history-taking system

The system consisted of patient- and HCP-facing tools (**Figure 1)**. The patient-facing tool was provided to recruited patients in the waiting room on a tablet computer (iPad→, Apple Inc.). The patient history taking tool used a cloud-based probabilistic reasoning engine combined with curated medical knowledge to ask the patient the optimal set of questions based on probable conditions and condition urgency. The Ada reasoning engine versions used in the study were 1.31.1 and 1.31.2, between August 3^rd^ - 21^st^ 2020. The assessment was performed as a question flow with options to confirm, deny or skip the question. The tool asked successively tailored questions about the respondent’s medical history, the main presenting complaint(s) and related attributes of their symptoms such as severity and time course.

**Figure 1.**
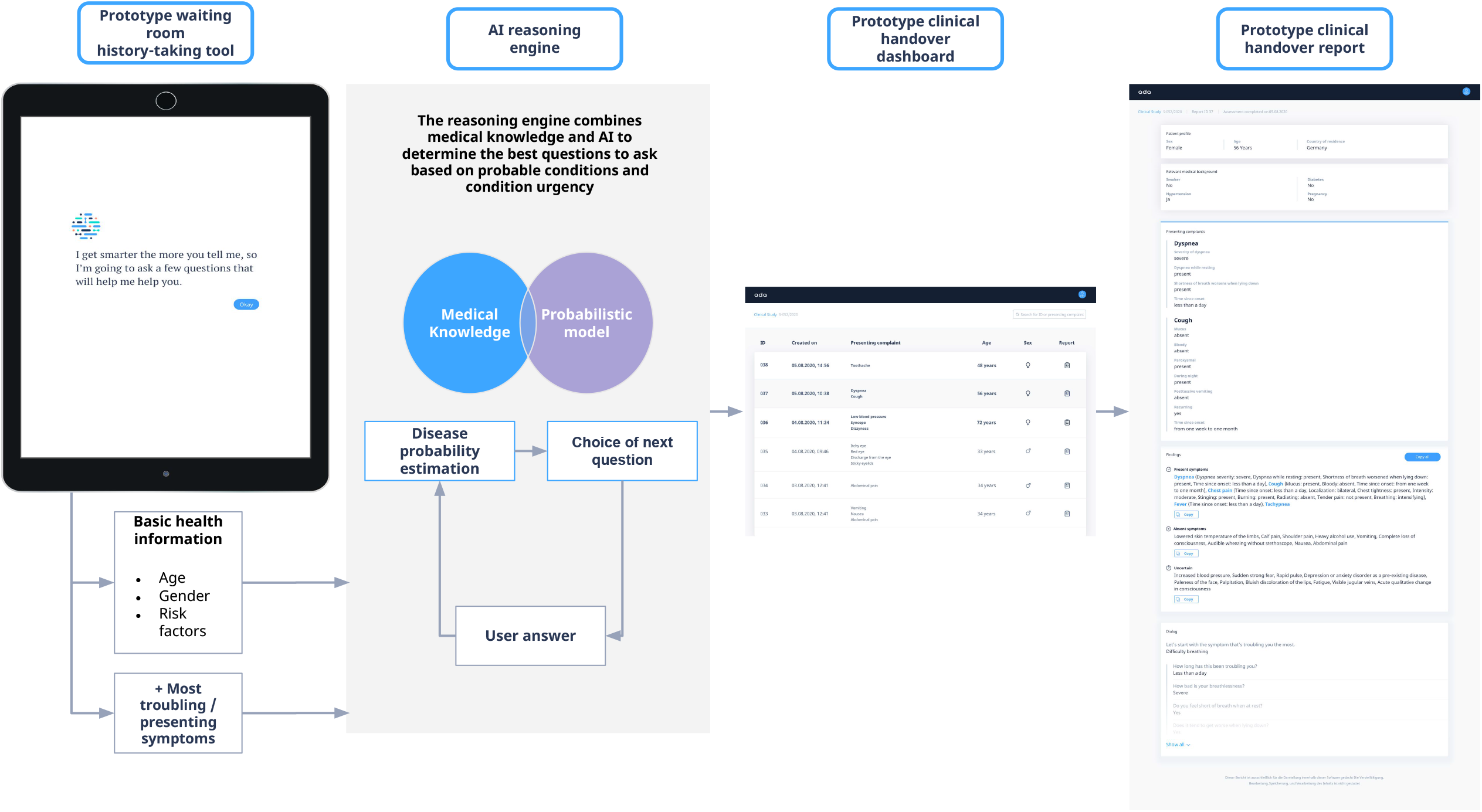
The prototype digital history/symptom-taking and handover system evaluated in this study, showing the interactions between the patient-facing and HCP-facing tools and describing how artificial intelligence reasoning engine functions to ask the patient, sequentially, the most relevant next question.

The HCP-facing tool provided a secure web-interface dashboard that listed all completed assessments to the ED clinical staff. The tool also provided a detailed handover report, designed to provide clinical information quickly and safely to HCPs. The handover report included the patient’s basic information (sex, year of birth), basic medical history information (smoker, hypertension, diabetes and pregnancy status), main presenting symptom(s), details of these symptoms, including the specific questions asked by the app, and answers provided by the patient.

### Design: study procedure

The procedure of the study is described in **Figure 2**.

**Figure 2.**
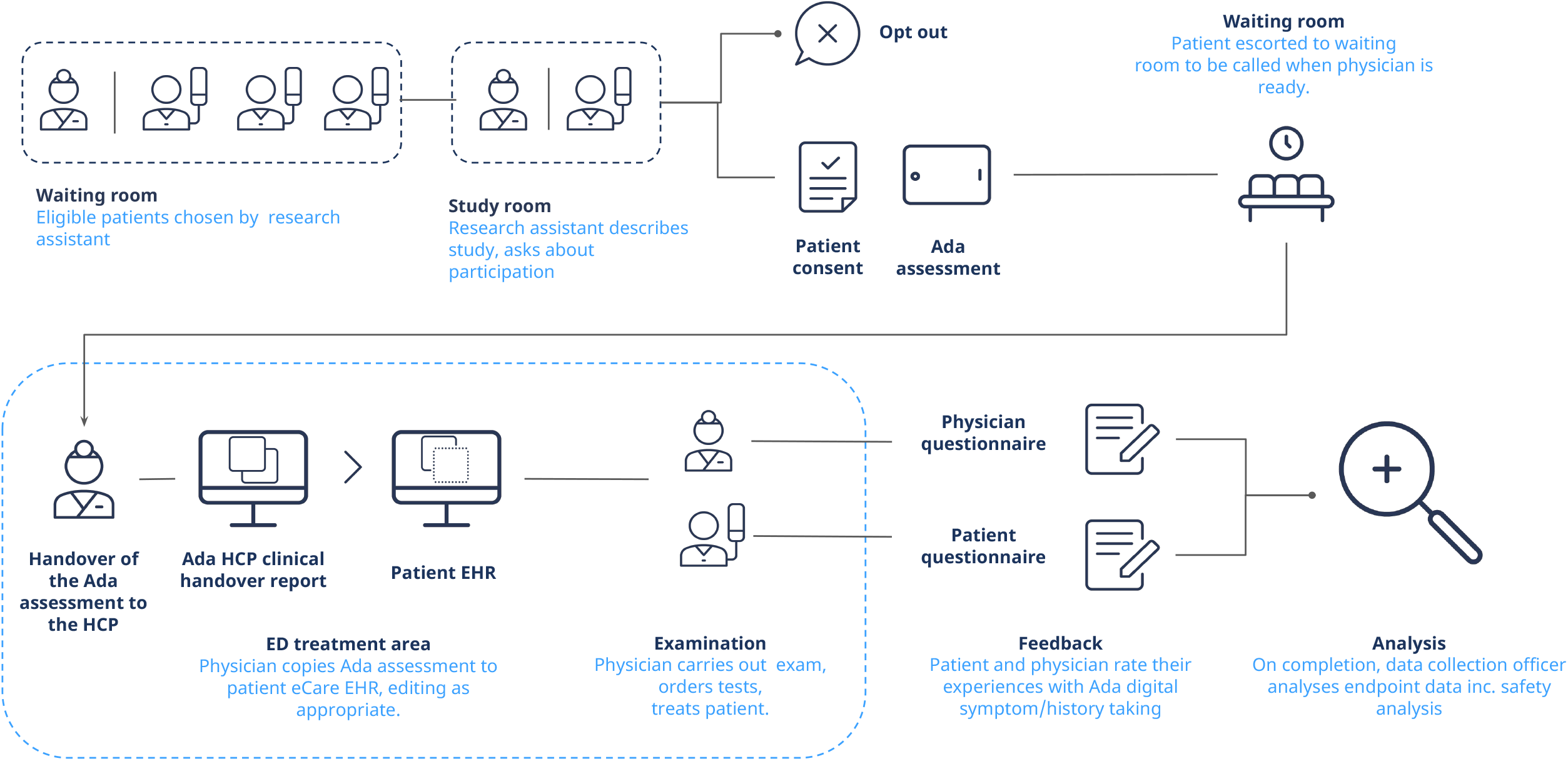
Overview of the study procedure.

#### Recruitment, inclusion and exclusion criteria

The inclusion criteria were: (i) all patients over 18 years old; (ii) attended the study sites when recruitment was being undertaken; (iii) willing and able to provide consent. Exclusion criteria were: (i) patients with severe injury/illness requiring immediate treatment, i.e. emergency severity index (ESI) levels 1-3; ^13^ (ii) patients with traumatic injury; (iii) patients incapable of completing a health assessment, e.g. due to illiteracy, mental impairment or inebriation or other incapacity; and, (iv) patients under 18 years old. The clinician research assistant asked all potentially eligible patients in the waiting room if they would be interested in participating in a study evaluating an app to record and pass their symptoms on to the ED doctors. Potentially eligible patients were informed that the study would not delay (or accelerate) their treatment at the ED. A single clinician research assistant carried out enrollment and consent.

#### Informed consent and study data management

If the patient agreed to be considered for inclusion, they were led to a separate private room adjacent to the ED where the nature, background and scope of the study were explained, and they were asked if they wanted to participate. If the patient consented the pseudo-anonymization procedure was followed. The patient’s name was recorded alongside the next-in-sequence study patient enrollment number on the study enrollment (disambiguation) record, which is the only link between the study ID (study patient enrollment number) and patient name, which was kept securely by the Principal Investigator. All quantitative and qualitative data was only accessible by the study team on secure systems. The patient’s enrollment number was entered into the patient-facing tool on the iPad and the tool then asked the patient to agree to the Terms and Conditions and Privacy Policy. The study team was familiar with data privacy regulations and committed to data protection principles. The study was approved by the local ethics committee at the University of Heidelberg (No. S-052-2020).

#### Procedure for patient symptom taking

The clinician research assistant answered any questions the patient had about the app and helped them to use it if requested, recording the degree of help provided. The symptom-taking handover report was not provided to the patient and was automatically made available to the ED physicians in the ED treatment area via the HCP-facing tool.

#### Training and study procedure for HCPs in the ED

All the ED nurses and physicians were made aware of the study in advance via a presentation of the study and a manual describing the system. Physicians were made aware of all patients who were enrolled in the study. The HCP logged onto the secure web interface using a secure ID to access the handover report.

### Study measurements

After examination by the ED physician, the patient and the ED physician completed separate questionnaires, with evaluation ratings of the tool on modified Likert scales. Nurses completed the same questionnaire as doctors when possible, however, it was recognized at the time of trial design, that there would not always be a nurse-patient interaction after triage in which the handover report is relevant.

### Study setting

The study was conducted in the ED of the Katharinenhospital Stuttgart which is an adult tertiary referral and major trauma hospital in South-Western Germany It provides interdisciplinary emergency treatment for 100-120 patients per day. The center adopts the First View Concept ^14^, in which an emergency registrar/consultant sees each patient in an interdisciplinary approach. The center has 23 treatment rooms with central monitoring, one resuscitation rooms, one wound and one plaster room. It uses the internationally recognized Emergency Severity Index (ESI) triage system to guide the treatment of emergency patients according to medical urgency ^13^.

### Sample size calculation

This study was designed for real-world tool optimization (in the action-oriented Phase I), followed by a preliminary observation assessment of the tool’s potential in the ED. It was also designed as guide to a later larger trial. The study sample size was determined by the requirement to evaluate the study safety criterion of identifying any adverse events of 5% using the approach of Viechtbauer et al.^15^: if a safety problem that exists with 5% population probability then the problem would be identified with 95% confidence in a sample size of 59 participants. The sample size calculation was for all patients recruited in both study phases, on the basis that no changes to the core AI assessment or core information in the handover report between versions would be carried out.

### Data analysis

The quantitative data were analyzed using standard Python (version 3.7.4) statistical modules (scipy module version 1.3.0) using descriptive statistics and the Mann-Whitney U test, a nonparametric test of statistical significance suitable for categorical data ^16^. For statistical significance testing, the value of α (here α = 0.05) was adjusted using the Bonferroni correction, α/*m*, where *m* is the number of questions evaluated for each group. For patients *m* = 6 and α_*corrected*_ = 0.0083 patients, for HCPs *m* = 4 and α_*corrected*_ = 0.0125 and for the degree of patient self-sufficiency *m* = 1 and α_*corrected*_ = 0.05.

## Results

### Recruitment

The total number of recruited patients was 81 (41 women, 40 men) with 45 patients in Phase I and 36 in Phase II (**Figure 3)**. Of the 81 recruited patients there were three ESI level 3 patients, 77 ESI level 4 patients and one ESI level 5 patient. A detailed description of the patient population is provided in **Table 1** and the full results for each patient are in **Supplementary Table 2**.

**Table 1.** Description of the sex, data-completeness and medical subdiscipline of the final main diagnosis stratified according to study phase. The number of patients in each group is expressed as n [the number of patients for whom a questionnaire exists]/N_total_ [the total number of recruited patients in this phase], followed by the percentage in brackets (%). ‡ - percent out of all patients recruited rather than % out of this study phase).

| Group by study phase | All patients | Phase I patients | Phase II patients |
| --- | --- | --- | --- |
| **Total: n/N<sub>total</sub> (%)** | 81/81 (100%) | 45/81‡(55.6%) | 36/81‡(44.4%) |
| **Age (mean, years)** | 38.7±15 | 36±15.9 | 42.1±13.1 |
| **Completed patient evaluation questionnaire** | 81/81 (100%) | 45/45 (100%) | 36/36 (100%) |
| **Completed physician evaluation questionnaire** | 78/81 (96.3%) | 43/45 (95.6%) | 35/36 (97.2%) |
| **Completed optional nurse evaluation questionnaire** | 10/81 (12.3%) | 1/45 (2.2%) | 9/36 (25%) |
| **Medical specialism classification (on basis of ED discharge diagnosis)** |  |  |  |
| **No Diagnosis Assigned** | 1/81 (1.2%) | 1/45 (2.2%) | 0/36 (0%) |
| **Orthopedics** | 12/81 (14.8%) | 9/45 (20%) | 3/36 (8.3%) |
| **Dermatology** | 5/81 (6.2%) | 5/45 (11.1%) | 0/36 (0%) |
| **Internal Medicine Including sub-specialisms** | 28/81 (34.6%) | 13/45 (11.1%) | 15/36 (41.7%) |
| **Internal Medicine Cardiovascular Disease** | 4/81 (4.9%) | 1/45 (2.2%) | 3/36 (8.3%) |
| **Internal Medicine Gastroenterology** | 4/81 (4.9%) | 1/45 (2.2%) | 3/36 (8.3%) |
| **Internal Medicine Nephrology** | 1/81 (1.2%) | 1/45 (2.2%) | 0/36 (0%) |
| **Internal Medicine Oncology** | 1/81 (1.2%) | 0/45 (0%) | 1/36 (2.8%) |
| **Internal Medicine Rheumatology** | 1/81 (1.2%) | 1/45 (2.2%) | 0/36 (0%) |
| **Internal Medicine With no subspecialism** | 17/81 (21%) | 9/45 (20%)") | 8/36 (22.2%) |
| **Neurology** | 20/81 (24.7%) | 8/45 (17.8%)") | 12/36 (33.3%) |
| **Obstetrics and Gynecology** | 1/81 (1.2%) | 1/45 (2.2%)") | 0/36 (0%) |
| **Psychiatry** | 1/81 (1.2%) | 1/45 (2.2%)") | 0/36 (0%) |
| **Surgery** | 10/81 (12.3%) | 6/45 (13.3%)") | 4/36 (11.1%) |
| **Ear, Nose and Throat (ENT)** | 3/81 (3.7%) | 1/45 (2.2%)") | 2/36 (5.6%) |

**Figure 3.**
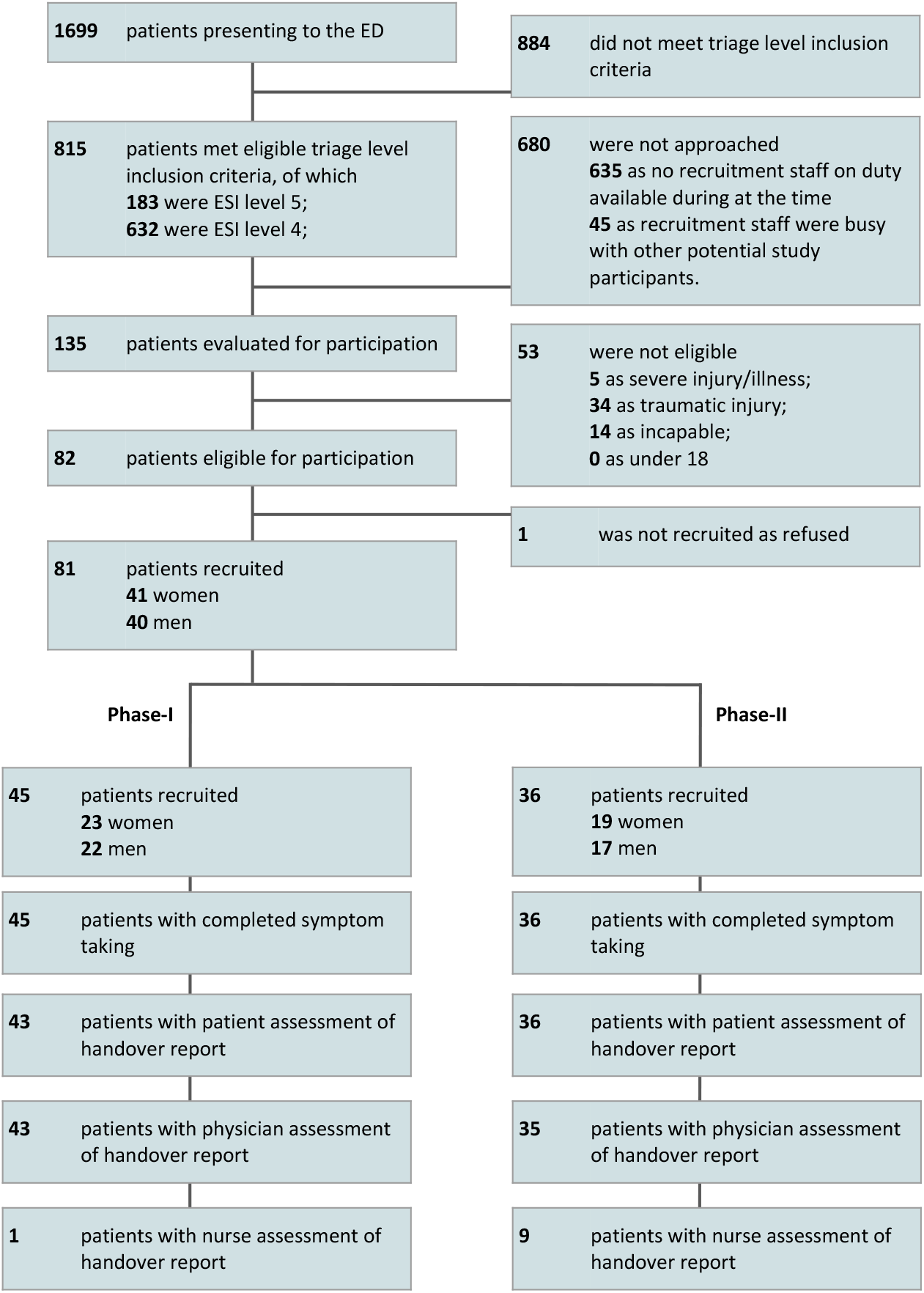
Participant Recruitment Flow

### Baseline Data

Patient populations in the two study phases were similar (**Figure 3** and **Table 1**): the mean (SD) age was 38.7±15.0 years for all patients, 36.0±15.9 for Phase I patients, and 42.1±13.1 for Phase II patients. The dominant medical classifications (by ED discharge diagnosis) were Orthopedics; Internal Medicine; Neurology and Surgery.

### Missing Data

Questionnaire completion rate was 100.0% for patients (of which 90.1% completed all questions), 96.3% for physicians and 12.3% for nurses (**Figure 3, Table 1 & 2**). Nurse questionnaires were only completed when they took part in with symptom/history information, and when ED busyness allowed. For all survey questions, the analysis approach was to report all data with respect to the number of responses to that survey question (i.e. with the denominator in analyses being lower where there was missing data).

### Safety Endpoint

A safety analysis was carried out for each patient by examining the handover report and the patient’s clinical history in the electronic health record, which were considered together, and alongside feedback from the treating HCPs, to determine if there was any evidence of inappropriate HCP guidance. The conclusion of this evaluation was that there were no cases of HCPs being guided in an unsafe manner by the handover report for the 81 recruited patients.

### Summary of changes made between the V1 and V2 systems

Three changes were made in total:

**1 - The addition of three free-text fields, where the patient can supply initial information on their medical history, current medications, and allergies**.

This change was requested by ED-physicians so that patients would have the opportunity to pass on information, in their own words, that was not always collected by the tool’s question flow. This was implemented in a manner that did not change the core AI-based symptom assessment. This change affected the information entered into the patient-facing tool, and the information presented on the HCP-facing tool.

**2 - Improve the user interface at the transition between patient information and symptom assessment**

A minor rephrasing was made to the patient-facing tool in response to a small number of patients who reported being confused by the initial wording.

**3 - Fix of a minor bug affecting the HCP-handover report**

A minor bug was removed which had resulted in a small number of handover reports not accurately displaying the transcript of questions that Ada asked the patient alongside the patient response.

The changes were anticipated to have a minor impact on the evaluation of the tool. The patients’ ratings of the tool (**Table 2**) improved by 9.5% and the physicians by 9.1% across all ratings with a significant improvement (p = .003, α_*corrected*_ = 0.0083) in the responses to the patient question ‘*(i) was the use of the tool and the answering of its questions interesting for you?’*. The improvements in nurse ratings of the system between Phase I and II are reported in **Table 2**, however, there were insufficient nurse assessments in Phase I to allow statistical significance testing.

**Table 2.**
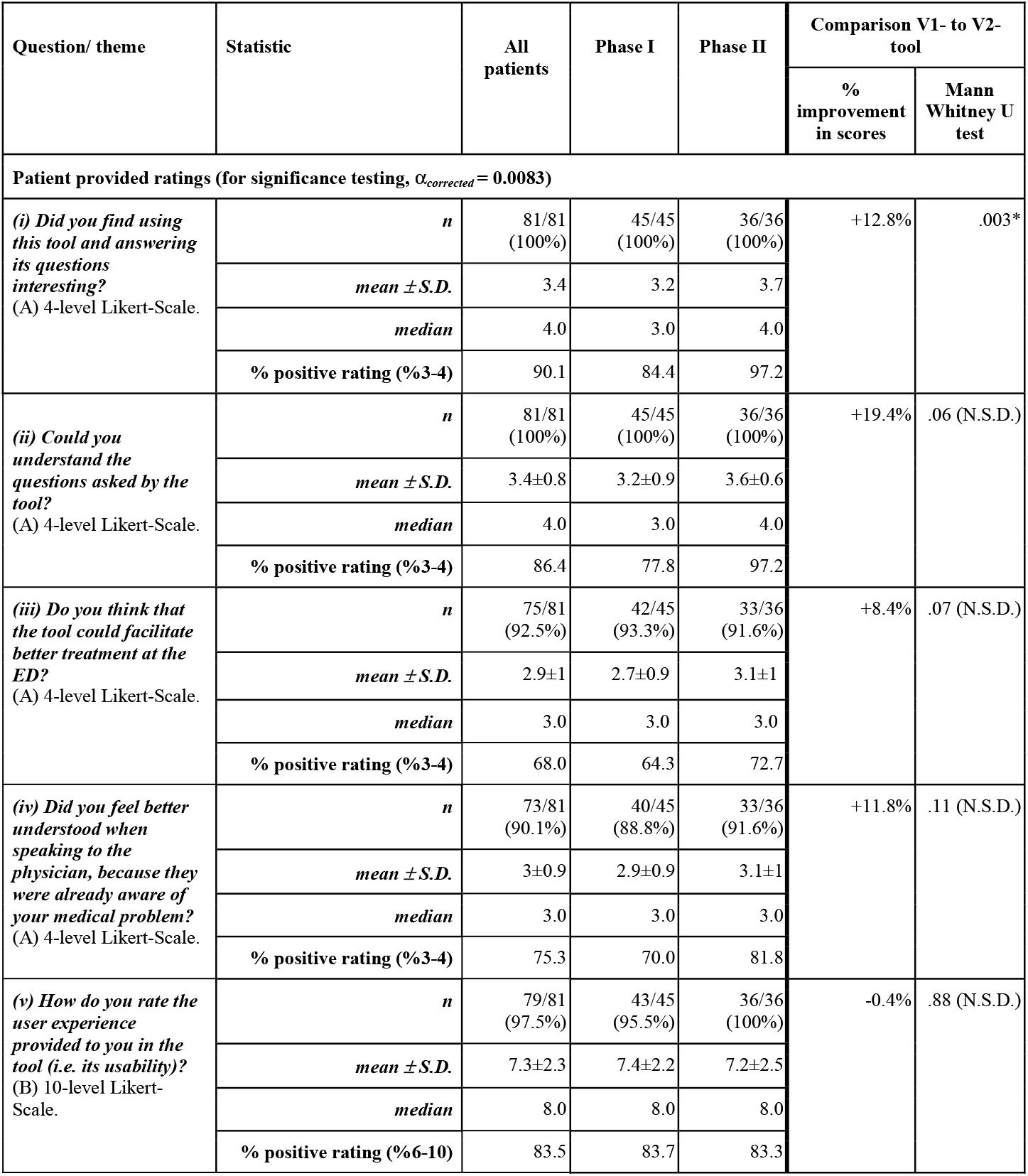
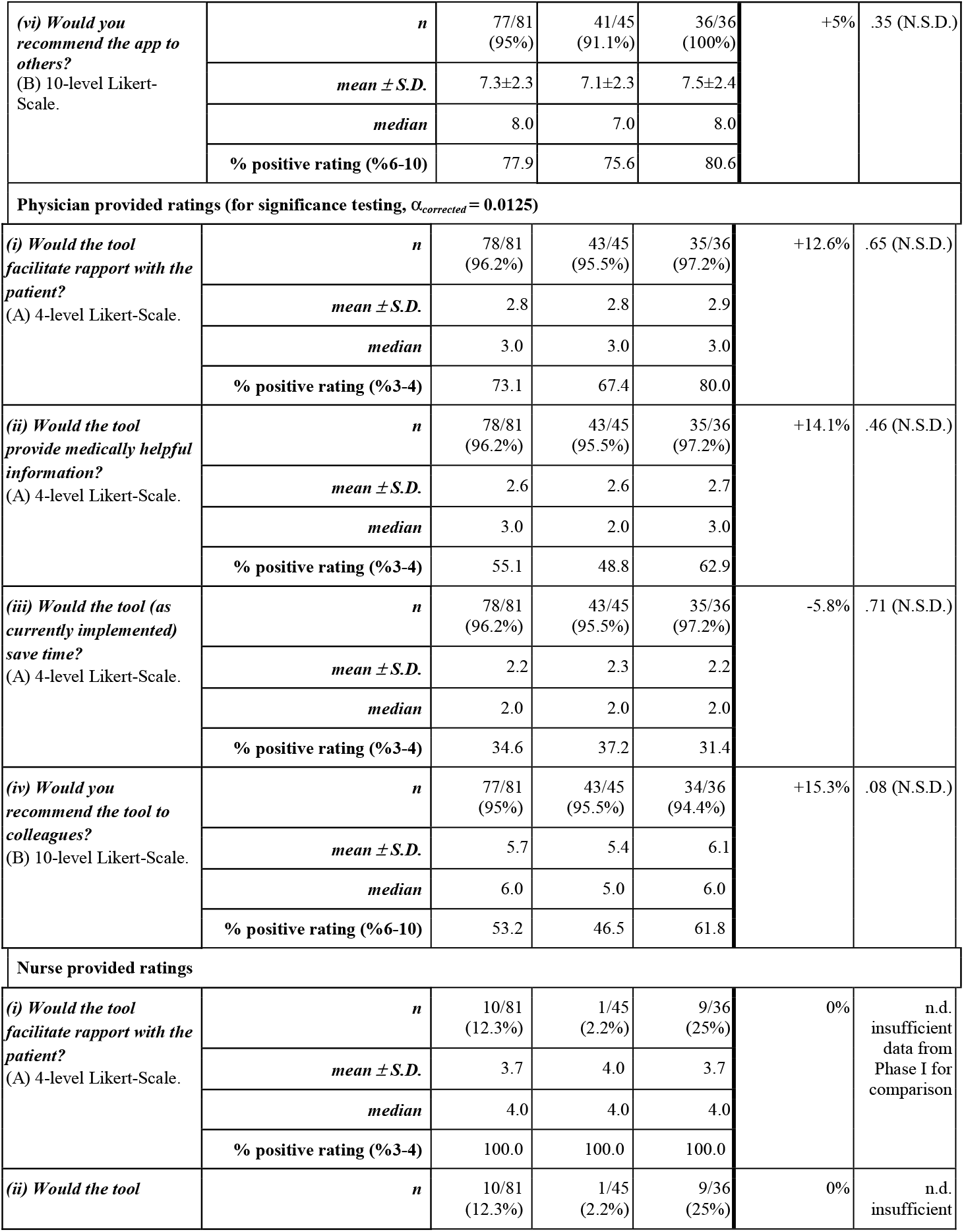
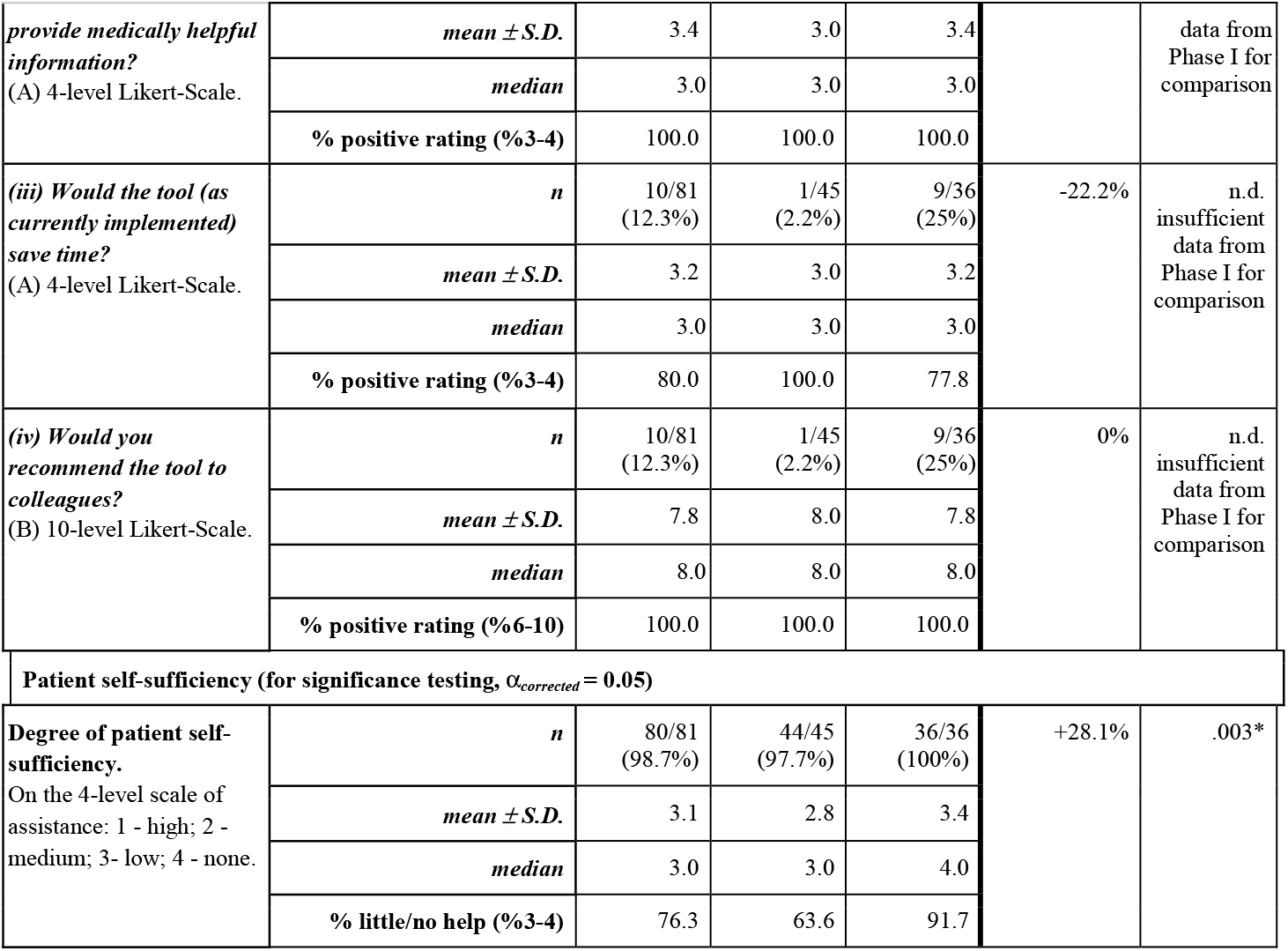
Summary of patient, physician and nurse ratings of the tool for Phase I, Phase II and the phases combined. Two modified Likert scales were used: 4-level Likert-Scale (A): *(1 - Strongly Disagree; 2 - Disagree; 3 - Agree; 4 - Strongly Agree)*, 10-level Likert-Scale (B): *(1 - Unlikely, to, 10 - Highly Likely)*. n.d.: not defined. Quality of evidence rating: 4 (*Oxford Centre of Evidence-based Medicine ratings of individual studies*). The mean, median and % positive ratings are calculated on the basis of the provided answers for each question. The number of responses is expressed as n [the number of responses to this question]/N_total_ [the total number of recruited patients in this phase], followed by the percentage in brackets (%).

### Patient, physician and nurse ratings of the tool

Patients were positive or highly positive (90.1%) about how interesting the tool was to use, its understandability (86.4%), its usability (83.5%), its ability to facilitate understanding and rapport with the HCPs (75.3%) and about recommending the tool to peers (83.5%) (**Table 2**). Likewise, 73.1% of physicians were positive or highly positive about the tool’s potential for facilitating understanding and rapport with patients.

Nurses were even more positive or highly positive about the tool’s potential for facilitating understanding and rapport with patients (100.0%), about the helpfulness of the medical information provided (100.0%), about the tool’s time saving potential (80.0%) and about recommending the tool to peers (100.0%).

### Subanalyses by medical specialism

A subanalysis by medical specialism of ED discharge diagnosis is shown in **Table 3** and described in detail in the **Supplementary Information**. This subanalysis showed there was lower understandability of the tool by Neurology patients, and these patients also gave lower ratings to the conversation facilitation provided by the tool. Physicians rated the level of helpful information, the time saving utility of the report and their likelihood to recommend the system more positively for Internal Medicine handovers than for handovers for all recruited patients or for other specialisms.

**Table 3.** Summary of patient, physician and nurse ratings of the tool, on a modified Likert scale, for the combined phases-I and II of the study according to medical subspecialism. Two modified Likert scales were used: 4-level Likert-Scale (A): *(1 - Strongly Disagree; 2 - Disagree; 3 - Agree; 4 - Strongly Agree)*, 10-level Likert-Scale (B): *(1 - Unlikely, to, 10 - Highly Likely)*. n.d.: not defined; Intern Med: Internal Medicine - Intern Med (no sub) - Internal Medicine (with no subspecialism); Neurol - Neurology; Ortho - Orthopedics; Surg - Surgery. Quality of evidence rating: 4 (*Oxford Centre of Evidence-based Medicine ratings of individual studies*). The number of responses is expressed as n [the number of responses to this question]/N_total_ [the total number of recruited patients in this phase], followed by the percentage in brackets (%).

| Question/ theme | Statistic | Intern Med | Intern Medi (no sub) | Neurol | Ortho | Surg |
| --- | --- | --- | --- | --- | --- | --- |
| **Patient provided ratings** |  |  |  |  |  |  |
| **(i) Was the use of the tool and the answering of its questions interesting for you?**<br>(A) 4-level Likert-Scale. | **n** | 24/81<br>(29.6%) | 17/81<br>(20.9%) | 20/81<br>(24.6%) | 5/81<br>(6.1%) | 10/81<br>(12.3%) |
| | **mean ± S.D.** | 3.3±0.7 | 3.3±0.7 | 3.6±0.6 | 3.8±0.4 | 3.6±0.5 |
|  | **median** | 3.0 | 3.0 | 4.0 | 4.0 | 4.0 |
|  | **% positive rating (%3-4)** | 83.3 | 88.2 | 95.0 | 100.0 | 100.0 |
| **(ii) Could you understand the questions asked by the tool?**<br>(A) 4-level Likert-Scale. | **n** | 24/81<br>(29.6%) | 17/81<br>(20.9%) | 20/81<br>(24.6%) | 5/81<br>(6.1%) | 10/81<br>(12.3%) |
| | **mean ± S.D.** | 3.5±0.7 | 3.5±0.5 | 3.1±0.9 | 3.6±0.5 | 3.4±0.7 |
|  | **median** | 4.0 | 4.0 | 3.0 | 4.0 | 3.5 |
|  | **% positive rating (%3-4)** | 91.7 | 100.0 | 75.0 | 100.0 | 90.0 |
| **(iii) Do you think that the tool could facilitate better treatment at the ED?**<br>(A) 4-level Likert-Scale. | **n** | 23/81<br>(28.3%) | 16/81<br>(19.7%) | 18/81<br>(22.2%) | 5/81<br>(6.1%) | 9/81<br>(11.1%) |
| | **mean ± S.D.** | 3±0.9 | 2.9±1 | 2.9±1 | 3.4±0.5 | 3±0.7 |
|  | **median** | 3.0 | 3.0 | 3.0 | 3.0 | 3.0 |
|  | **% positive rating (%3-4)** | 69.6 | 62.5 | 66.7 | 100.0 | 77.8 |
| **(iv) Did you feel better understood when speaking to the physician, because they were already aware of your medical problem?**<br>(A) 4-level Likert-Scale. | **n** | 22/81<br>(27.1%) | 15/81<br>(18.5%) | 18/81<br>(22.2%) | 5/81<br>(6.1%) | 9/81<br>(11.1%) |
| | **mean ± S.D.** | 3.3±0.7 | 3.2±0.7 | 2.7±1 | 3.6±0.5 | 3.4±0.5 |
|  | **median** | 3.0 | 3.0 | 3.0 | 4.0 | 3.0 |
|  | **% positive rating (%3-4)** | 86.4 | 86.7 | 61.1 | 100.0 | 100.0 |
| **(v) How do you rate the user experience provided to you in the tool (i.e. its usability)?**<br>(B) 10-level Likert-Scale. | **n** | 24/81<br>(29.6%) | 17/81<br>(20.9%) | 20/81<br>(24.6%) | 5/81<br>(6.1%) | 10/81<br>(12.3%) |
| | **mean ± S.D.** | 7.1±2.2 | 7.2±2.2 | 6.8±2.6 | 8.2±1.9 | 8.4±0.8 |
|  | **median** | 7.5 | 8.0 | 7.5 | 9.0 | 8.0 |
|  | **% positive rating (%6-10)** | 79.2 | 82.4 | 80.0 | 80.0 | 100.0 |
| (vi) *Would you recommend the app to others?*<br>(B) 10-level Likert-Scale. | *n* | 23/81<br>(28.3%) | 16/81<br>(19.7%) | 20/81<br>(24.6%) | 5/81<br>(6.1%) | 10/81<br>(12.3%) |
|  | *mean ± S.D.* | 7±2.4 | 6.9±2.5 | 7.1±2.4 | 8.2±1.9 | 8.4±1.4 |
|  | *median* | 7.0 | 7.0 | 7.5 | 9.0 | 8.5 |
|  | % positive rating (%6-10) | 73.9 | 75.0 | 70.0 | 80.0 | 100.0 |
| **Physician provided ratings** |  |  |  |  |  |  |
| (i) *Would the tool facilitate rapport with the patient?*<br>(A) 4-level Likert-Scale. | *n* | 27/81<br>(33.3%) | 16/81<br>(19.7%) | 20/81<br>(24.6%) | 12/81<br>(14.8%) | 9/81<br>(11.1%) |
|  | *mean ± S.D.* | 2.9±0.8 | 2.8±0.9 | 2.8±0.8 | 2.5±1 | 3.4±0.7 |
|  | *median* | 3.0 | 3.0 | 3.0 | 2.5 | 4.0 |
|  | % positive rating (%3-4) | 81.5 | 68.8 | 75.0 | 50.0 | 88.9 |
| (ii) *Would the tool provide medically helpful information?*<br>(A) 4-level Likert-Scale. | *n* | 27/81<br>(33.3%) | 16/81<br>(19.7%) | 20/81<br>(24.6%) | 12/81<br>(14.8%) | 9/81<br>(11.1%) |
|  | *mean ± S.D.* | 2.7±0.7 | 2.6±0.9 | 2.7±0.8 | 2.5±1 | 3±0.9 |
|  | *median* | 3.0 | 3.0 | 3.0 | 2.0 | 3.0 |
|  | % positive rating (%3-4) | 66.7 | 62.5 | 60.0 | 41.7 | 66.7 |
| (iii) *Would the tool (as currently implemented) save time?*<br>(A) 4-level Likert-Scale. | *n* | 27/81<br>(33.3%) | 16/81<br>(19.7%) | 20/81<br>(24.6%) | 12/81<br>(14.8%) | 9/81<br>(11.1%) |
|  | *mean ± S.D.* | 2.3±0.7 | 2.4±0.8 | 2.3±0.9 | 1.9±1 | 2.8±1.1 |
|  | *median* | 2.0 | 2.5 | 2.0 | 2.0 | 3.0 |
|  | % positive rating (%3-4) | 40.7 | 50.0 | 30.0 | 25.0 | 55.6 |
| (iv) *Would you recommend the tool to colleagues?*<br>(B) 10-level Likert-Scale. | *n* | 27/81<br>(33.3%) | 16/81<br>(19.7%) | 19/81<br>(23.4%) | 12/81<br>(14.8%) | 9/81<br>(11.1%) |
|  | *mean ± S.D.* | 6.2±1.7 | 6.3±2.1 | 5.6±1.8 | 5.2±2.2 | 6±2.3 |
|  | *median* | 6.0 | 6.5 | 6.0 | 5.0 | 6.0 |
|  | % positive rating (%6-10) | 70.4 | 68.8 | 52.6 | 41.7 | 100.0 |
| **Patient self-sufficiency** |  |  |  |  |  |  |
| **Degree of patient self-sufficiency.**<br>On the 4-level scale of assistance: 1 - high;<br>2 - medium; 3- low; 4 - none. | *n* | 27/81<br>(33.3%) | 16/81<br>(19.7%) | 20/81<br>(24.6%) | 12/81<br>(14.8%) | 10/81<br>(12.3%) |
|  | *mean ± S.D.* | 3±1 | 3.4±0.6 | 3.2±1 | 3.1±0.9 | 3.3±0.8 |
|  | *median* | 3.0 | 3.0 | 3.0 | 3.0 | 3.5 |
|  | % little/no help (%3-4) | 77.8 | 93.8 | 80.0 | 66.7 | 80.0 |

### Degree of patient assistance provided

Across both phases, most patients were able to use the tool with little or no assistance (76.3%). This measure improved significantly between Phase I (63.6%_ and II (91.7%, p = .003, α_*corrected*_ = 0.05).

### Variability of physicians’ perceptions

Patient symptom and history data handover was evaluated for 96.3% (78/81) of the patients by 24 different ED physicians. Qualitative interviews with physicians in Phase I of the study revealed a number of physicians who were exceptionally enthusiastic about the performance of the system, i.e. an ‘early adopter’ mentality. The quantitative analysis of the distribution of Likert scores (see **Supplementary Figure 1**) supported these qualitative findings, in that there is a skewed distribution of physician scores, with two physicians (one of whom evaluated handover reports for 6 patients), providing highly positive evaluations of the tool.

## Discussion

### Overall results discussion

This study used methodologies that have been adopted in other ED quality improvement evaluations of new digital technologies^10,17^, to evaluate a novel AI-based symptom/history taking and handover system, by means of patient and HCP questionnaires. The performance of the system was evaluated in two principle areas: (i) the facilitation of bidirectional patient-to-clinician conversation and rapport building; and, (ii) the potential to save HCP time through symptom collection and documentation facilitation.

There was a strongly positive rating by patients, physicians and nurses of the tool as an aid in patient-to-clinician conversation, communication and rapport building. The proportion of physicians positive/highly positive about recommending the system to their peers (53.2%) was not as high as it was for patients and nurses (77.9% and 100.0% respectively). Overall, physicians had mixed views on the degree of helpful clinical information and time saving potential of the clinical handover report. Underlying the mixed results in the physician ratings (**Supplementary Table 2**), were a large number of patients for whom the ED physicians described the presence of a ‘visual diagnosis’ - i.e. one that was immediately apparent by simply looking at a patient, and for whom patient-directed symptom taking, in whatever form it is designed, is unlikely to save history-taking and recording time. For some other patients, the tool did not adequately draw together and summarize a highly complex, sometimes multimorbid medical presentation in a manner that made it useful for physicians. Both qualitative findings and quantitative analysis revealed a subset of highly enthusiastic early-adopter physicians, as have been recognized in other studies ^18^, who were highly positive about the history and symptom taking information provided by the tool and for its potential to improve rapport, to provide helpful clinical information and to save ED time.

The relatively lower understandability of the tool by Neurology patients, and the lower rating of its conversation facilitation, may reflect effects of neurological symptoms on their use of the tool. Our interpretation is that the positive/highly positive evaluations from physicians for Internal Medicine handovers is likely due to the system’s AI reasoning engine being better at directing the question flow for conditions which have many subtle interlinked clinical symptoms.

One aim for the development of a waiting room patient history taking - and HCP handover tool is that it be easy to use without assistance. Many patients visit only once, so they would have little opportunity to learn to use the tool over time. In this study, patient assistance was provided where needed by the recruiting physician (JSB) to maximize data collection for the action-orientated tool improvement and so that the ED-physician/nurse facing interface’s performance could be assessed with meaningful clinical information. Most (76.3%) patients required little or no help with the use of the tool, and in some subspecialisms, patients were highly independent using the tool (93.8% in Internal Medicine with no subspecialism), whereas 66.7% of Orthopedics patients required little or no help. A priority in the future development of the prototype will be to adapt the reasoning engine and the user interface to minimize the level of help required for all patient groups, and this may involve making the tool available in the patient’s primary language instead of in the national language only.

The action-orientated approach with two study phases provided an opportunity within a clinical trial framework to rapidly modify a prototype tool’s design, based on feedback from patient and physician evaluations, and to further evaluate the modified tool. Comparison of tool ratings showed statistically significant improvements in performance between the V1 and V2 tools in two evaluation categories: (i) *how engaging/interesting patients found the tool to use*; and, (ii) *the degree of self-sufficiency of the patients in using the tool*. These improvements can be explained through improvements made in tool usability and in functionality added to the tool to capture additional information about their symptoms and history. Many useful insights on tool performance and usability were obtained in the study. Only a relatively small number of optimizations could be executed between the V1 and V2 tools, due to the need to prioritize in-study changes to those that could be completed within the possible study recruitment period. The remaining insights, including those on the V2 tool will be used in later prototype development and optimization.

There are no previous studies describing similar digital history/symptom-taking and handover systems in the literature. Arora et al. (2014) ^19^ explored patient impressions and satisfaction of a self-administered, automated medical history-taking device in the ED, and they also reported high levels of patient enthusiasm in the evaluated tool and for the potential for rapport-building with HCPs. However, handover to HCPs was not carried out in their study, nor did the tool have a system to enable handover. A number of studies ^9,20,21^ have reported apps for self-assessment of urgency/triage in the general practice and acute primary care setting, but evaluation of the handover of history and symptom information to HCPs was not addressed.

### Clinical and Policy Implications and Implications for Future Research

It is recognized that empathy and rapport can be lacking in EDs, both of which are important for staff stress levels and satisfaction and for patient outcomes and empowerment ^1,5,6,22,23^. Although a digital tool can never be a complete solution to improve human-to-human empathy, this study has nonetheless shown patient and HCP enthusiasm for the rapport building and conversation facilitation enabled through such tools. This study evaluated a prototype, and optimizations of this tool are required, based on the insights from this study, to facilitate patient-HCP conversations and optimize rapport in a manner which is complete (i.e. clinically helpful information for every patient), streamlined and seamless (i.e. no additional tasks or systems for clinicians). This will then be followed by clinical trial evaluation of a completed tool.

Although the results of this study were definitive on the potential for rapport and conversation facilitation in the ED, they were equivocal on the potential for the tool, as it is currently implemented, to save clinician time, and clinicians could identify time saving only in some types of patient presentations (Internal Medicine, Surgery). Nurses were more positive about the general potential of the tool for clinician time saving. It is recognized that further development of the prototype is required if the aim of the completed approved regulatory product is to deliver definitive physician time savings for all patients. The role of a patient symptom and history taking tool, is not only potentially important in saving clinician time, but also in contributing to documentation accuracy and completeness, as it is known that medical performance reduces with stress and over-stretching in the ED, and is likely to result in greater error making, including in documentation ^4,5^. It was not possible to measure the contribution of the tool to documentation completeness and accuracy in this study.

### Limitations

This was a single site study in Germany and the findings may not be generalizable to other facilities or to other countries. The patient and physician evaluations were not influenced by selection bias, however, as the nurse evaluations were completed on a voluntary basis the influence of selection bias cannot be excluded and 90% of the nurse evaluations relate to study Phase II. This study explored tool use in the German language and in the German setting only, and the sample size was relatively small sample given resource constraints within the ED. The study did not include pediatric patients. Infection control with patients using a tablet in the ED could be challenging. This study explored the early phase after prototype system implementation, training and first experience of its application by users. Although minimally relevant for ED patients, the skill and speed of ED physician and nurse use of the new tool, and their perception of its performance is likely to change over time. Despite limitations, our approach allowed us to investigate implementation at this single site in considerable depth.

## Conclusions

The tool improved rapport between patient and clinician and improved patient-clinician communication. Notably, patients felt better understood, and the tool had utility in symptom gathering and nurses perceived it as having workflow benefit through time saving. Some physicians were enthusiastic about the potential to improve patient interaction and about the tool’s benefits in symptom and history taking. Results regarding time saving in the ED were equivocal, but there was potential for time saving in some medical subspecialisms e.g. in Internal Medicine and in Surgery. The insights from this study will be used for further prototyping and research to extend the range of patients for which the tool can provide helpful conversation support for patients and time savings for nurses, and iterate on the symptom and history collection capabilities to also provide more consistent time savings for clinicians. The tool is based upon an existing and regularly maintained medical reasoning engine, and therefore is a sustainable technological approach for symptom and history taking, and is readily adaptable to other related settings in which patient self-symptom and history taking and conversation support are relevant, including the at-home setting, the primary care setting and the specialist clinic setting (e.g. specialist rare diseases clinics).

## Data Availability

All data is included in the manuscript.

## Article Information

### Author Affiliations

Department for interdisciplinary Acute, Emergency and Intensive Medicine, (DIANI), Katharinenhospital, Klinikum Stuttgart, Kriegsbergstraße 60, 70174 Stuttgart, Germany; Ada Health GmbH, Karl-Liebknecht-Str. 1, 10178 Berlin, Germany.

### Author Contributions

Dr Scheder-Bieschin had full access to all of the data in the study and takes responsibility for the integrity of the data and the accuracy of the data analysis.

*Concept and design:* Scheder-Bieschin, Blümke, de Buijzer, Ondresik, Schilling, Gilbert.

*Coordination of the study in the emergency department:* Scheder-Bieschin, Schilling. *Acquisition, analysis, or interpretation of data:* Scheder-Bieschin, Blümke, Echterdiek, Nacsa, Ott, Paul, Schilling, Schmitt, Wicks, Gilbert.

*Drafting of the manuscript:* Scheder-Bieschin, Nacsa, Ondresik, Schmitt, Wicks, Gilbert.

*Statistical analysis:* Scheder-Bieschin, Schilling, Gilbert. *Administrative, technical, or material support:* Nacsa, Ondresik. *Supervision:* Scheder-Bieschin, Schilling, Gilbert.

### Data Sharing Statement

All data relevant to the study are included in the article or uploaded as supplementary information.

### Additional Contributions

None

