## Supplemental Material for "Improving emergency department patient-doctor conversation through an artificial intelligence symptom taking tool: an action-oriented design pilot study"

### Results Supplement - Subanalyses by medical specialism

A subanalysis by medical specialism of ED discharge diagnosis was carried out for all medical specialisms for which there were 10% or more of the patients included (Internal Medicine, Internal Medicine (with no subspecialism), Neurology, Orthopedics, and Surgery) is presented in (**Data Supplement Table 2**). Patient self-sufficiency in the use of the tool was relatively lower in Neurology (66.7% requiring little/no help) (compared to 76.3% for all patients).

In terms of patient evaluations, the subanalysis groups had similar ratings to the all-patient analysis, except in the following themes, were there was greater than 10% difference between the patients in the specialism and the full study population: (i) 75.0% of Neurology patients (compared to 86.4% of all patients) gave a positive/strongly positive evaluation for the understandability of tool questions; (ii) 100.0% of Orthopedics patients (compared to 68.0% of all patients) gave a positive/strongly positive evaluation for their view of the potential for the tool to facilitate better treatment in the ED; (iii) 61.1% of Neurology patients (compared to 75.3% of all patients) gave a positive/strongly positive evaluation for the tools ability to increase their being understood when speaking to the physician; (iv) 100.0% of Surgery patients (compared to 83.5% of all patients) gave a positive/strongly positive evaluation for the tools usability; and, (v) 100.0% of Surgery patients (compared to 77.9% of all patients) would recommend the tool to fellow patients.

For physician evaluations, there were similar ratings in the subanalyses to the all-patient analysis, except in the following themes: (i) 66.7% of Internal Medicine (all) and 66.7% of Surgery patient handovers (compared to 55.1% of all handovers) gave a positive/strongly positive evaluation for the provision of medically helpful information; (ii) 50.0% of Internal Medicine (with no subspecialism) and 55.6% of Surgery patient handovers (compared to 34.6% of all handovers) gave a positive/strongly positive evaluation for the potential for the tool to save time for the physician; (iii) 70.4% of Internal Medicine (all), 68.8% of Internal Medicine (with no subspecialism) and 100.0% of Surgery patient handovers (compared to 53.2% of all handovers) would positively/strongly positively recommend the tool to other physicians.

**Supplementary Table 2** The data for each patient and for each phase of the study.

| **ID** | **Study phase** | **Discharge diagnosis** | **Patient provided ratings** | | | | | | **Physician provided ratings** | | | | | | | | **Self-sufficiency** |
| --- | --- | --- | --- | --- | --- | --- | --- | --- | --- | --- | --- | --- | --- | --- | --- | --- | --- |
|  |  |  | **(i) Did you find using this tool and answering its questions interesting?**  **(A) 4-level Likert-Scale.** | **(ii) Could you understand the questions asked by the tool?**  **(A) 4-level Likert-Scale.** | **(iii) Do you think that the tool could facilitate better treatment at the ED?**  **(A) 4-level Likert-Scale.** | **(iv) Did you feel better understood when speaking to the physician, because they were already aware of your medical problem?**  **(A) 4-level Likert-Scale.** | **(v) How do you rate the user experience provided to you in the tool (i.e. its usability)?**  **(B) 10-level Likert-Scale.** | **(vi) Would you recommend the app to others?**  **(B) 10-level Likert-Scale.** | **(i) Would the tool facilitate rapport with the patient?**  **(A) 4-level Likert-Scale.** | | **(ii) Would the tool provide medically helpful information?**  **(A) 4-level Likert-Scale.** | | **(iii) Would the tool (as currently implemented) save time?**  **(A) 4-level Likert-Scale.** | | **(iv) Would you recommend the tool to colleagues?**  **(B) 10-level Likert-Scale.** | | ***Degree of patient self-sufficiency.***  ***On the 4-level scale of assistance: 1 - high; 2 - medium; 3- low; 4 - none.*** |
|  |  |  | **Patient** | **Patient** | **Patient** | **Patient** | **Patient** | **Patient** | **Dr** | **Nurse** | **Dr** | **Nurse** | **Dr** | **Nurse** | **Dr** | **Nurse** | **Researcher** |
| 001 | Phase I | Internal Medicine | 3 | 4 | 1 | 2 | 9 | - | 1 | 4 | 1 | 3 | 1 | 3 | 1 | 8 | 4 |
| 002 | Phase I | Orthopedics | 4 | 4 | 4 | 4 | 9 | 9 | 2 | - | 2 | - | 1 | - | 5 | - | 2 |
| 003 | Phase I | Surgery | 4 | 4 | 4 | 4 | 8 | 6 | 2 | - | 2 | - | 2 | - | 3 | - | 2 |
| 004 | Phase I | Orthopedics | 3 | 4 | 3 | 3 | 8 | 10 | 3 | - | 4 | - | 3 | - | 7 | - | 2 |
| 005 | Phase I | Orthopedics | 4 | 3 | 4 | 4 | 5 | 5 | 3 | - | 2 | - | 1 | - | 5 | - | 2 |
| 006 | Phase I | IM-Rheumatology | 4 | 4 | 4 | 4 | 7 | 7 | 4 | - | 3 | - | 3 | - | 7 | - | 1 |
| 007 | Phase I | Orthopedics | 4 | 4 | 3 | 3 | 10 | 9 | 3 | - | 3 | - | 2 | - | 6 | - | 3 |
| 008 | Phase I | General Surgery | 4 | 4 | 3 | 3 | 8 | 10 | 3 | - | 3 | - | 3 | - | 5 | - | 3 |
| 009 | Phase I | Internal Medicine | 4 | 3 | 3 | 3 | 5 | 5 | 4 | - | 4 | - | 4 | - | 9 | - | 2 |
| 010 | Phase I | General Surgery | 3 | 3 | 2 | 3 | 7 | 7 | 4 | - | 4 | - | 4 | - | 9 | - | 3 |
| 011 | Phase I | General Surgery | 3 | 4 | 3 | 4 | 10 | 10 | 4 | - | 3 | - | 4 | - | 9 | - | 2 |
| 012 | Phase I | Orthopedics | 4 | 3 | 3 | 4 | 9 | 8 | 4 | - | 4 | - | 4 | - | 9 | - | 4 |
| 013 | Phase I | Psychiatry | 2 | 1 | 2 | 1 | 2 | 2 | 3 | - | 2 | - | 1 | - | 6 | - | 1 |
| 014 | Phase I | ENT | 3 | 4 | 1 | - | - | - | 4 | - | 4 | - | 4 | - | 10 | - | 3 |
| 015 | Phase I | Neurology | 4 | 2 | 3 | 3 | 8 | 5 | 4 | - | 4 | - | 4 | - | 9 | - | 3 |
| 016 | Phase I | Neurology | 2 | 1 | 1 | 2 | 4 | 3 | 1 | - | 1 | - | 1 | - | 2 | - | 3 |
| 017 | Phase I | Neurology | 3 | 2 | 3 | 2 | 6 | 6 | 3 | - | 3 | - | 2 | - | 7 | - | 1 |
| 018 | Phase I | Neurology | 3 | 3 | 2 | 2 | 1 | 4 | 3 | - | 2 | - | 2 | - | 5 | - | 1 |
| 019 | Phase I | IM-Nephrology | 2 | 2 | 2 | 2 | 3 | 3 | 3 | - | 3 | - | 2 | - | 6 | - | 1 |
| 020 | Phase I | Internal Medicine | 2 | 3 | 2 | 3 | 7 | 6 | 4 | - | 3 | - | 3 | - | 7 | - | 4 |
| 021 | Phase I | IM-Cardiology | 4 | 4 | 3 | 3 | 8 | 10 | 3 | - | 3 | - | 2 | - | 5 | - | 3 |
| 022 | Phase I | Dermatology | 2 | 1 | 1 | 2 | - | - | 2 | - | 2 | - | 2 | - | 4 | - | 2 |
| 023 | Phase I | Orthopedics | 4 | 3 | 3 | 3 | 8 | 8 | 1 | - | 1 | - | 1 | - | 2 | - | 2 |
| 024 | Phase I | Orthopedics | 4 | 4 | 4 | 4 | 10 | 6 | 1 | - | 1 | - | 1 | - | 2 | - | 3 |
| 025 | Phase I | IM-Gastroenterology | 2 | 2 | 3 | 3 | 8 | 8 | 3 | - | 3 | - | 2 | - | 5 | - | 1 |
| 026 | Phase I | Internal Medicine | 2 | 4 | 3 | 3 | 10 | 8 | 3 | - | 4 | - | 3 | - | 6 | - | 4 |
| 027 | Phase I | Internal Medicine | 3 | 3 | 4 | 4 | 6 | 5 | 3 | - | 2 | - | 2 | - | 4 | - | 3 |
| 028 | Phase I | Dermatology | 3 | 4 | 2 | 2 | 10 | - | 2 | - | 2 | - | 1 | - | 4 | - | 3 |
| 029 | Phase I | Neurology | 3 | 2 | 3 | 2 | 6 | 6 | 2 | - | 2 | - | 2 | - | 3 | - | 2 |
| 030 | Phase I | No diagnosis assigned | 4 | 4 | - | - | 6 | 5 | - | - | - | - | - | - | - | - | 3 |
| 031 | Phase I | Dermatology | 3 | 2 | 3 | 3 | 8 | 6 | 2 | - | 2 | - | 2 | - | 3 | - | 4 |
| 032 | Phase I | Neurology | 4 | 4 | 2 | 1 | 10 | 10 | 3 | - | 2 | - | 2 | - | 8 | - | 4 |
| 033 | Phase I | Dermatology | 4 | 4 | 2 | 2 | 8 | 6 | 3 | - | 2 | - | 1 | - | 4 | - | 4 |
| 034 | Phase I | Internal Medicine | 3 | 4 | - | - | 8 | 3 | - | - | - | - | - | - | - | - | - |
| 035 | Phase I | Neurology | 3 | 4 | - | - | 10 | 10 | 3 | - | 4 | - | 3 | - | 6 | - | 4 |
| 036 | Phase I | Neurology | 4 | 3 | 3 | 3 | 8 | 10 | 3 | - | 3 | - | 2 | - | 4 | - | 4 |
| 037 | Phase I | General Surgery | 4 | 3 | 4 | 4 | 8 | 9 | 3 | - | 2 | - | 2 | - | 6 | - | 4 |
| 038 | Phase I | Orthopedics | 3 | 4 | 3 | 3 | 7 | 8 | 2 | - | 1 | - | 1 | - | 4 | - | 3 |
| 039 | Phase I | General Surgery | 3 | 2 | 3 | 3 | 8 | 8 | 4 | - | 2 | - | 3 | - | 6 | - | 4 |
| 040 | Phase I | Orthopedics | 3 | 4 | 3 | 3 | 8 | 8 | 2 | - | 2 | - | 2 | - | 3 | - | 4 |
| 041 | Phase I | Dermatology | 4 | 4 | 1 | 1 | 10 | 10 | 2 | - | 2 | - | 2 | - | 3 | - | 3 |
| 042 | Phase I | Gynecology | 3 | 3 | 3 | 3 | 6 | 6 | 3 | - | 3 | - | 3 | - | 5 | - | 2 |
| 043 | Phase I | Internal Medicine | 3 | 3 | 3 | 3 | 7 | 7 | 3 | - | 3 | - | 3 | - | 7 | - | 4 |
| 044 | Phase I | Internal Medicine | 3 | 3 | 2 | - | 5 | 9 | 3 | - | 3 | - | 3 | - | 6 | - | 3 |
| 045 | Phase I | Internal Medicine | 3 | 3 | 2 | 3 | 9 | 9 | 2 | - | 2 | - | 2 | - | 5 | - | 3 |
| 046 | Phase II | IM-Gastroenterology | 3 | 3 | 3 | 3 | 7 | 8 | 3 | - | 2 | - | 2 | - | 6 | - | 2 |
| 047 | Phase II | Internal Medicine | 4 | 4 | 4 | 4 | 9 | 9 | 3 | - | 2 | - | 2 | - | 5 | - | 3 |
| 048 | Phase II | IM-Cardiology | 4 | 4 | 4 | 4 | 2 | 2 | 3 | - | 2 | - | 2 | - | 7 | - | 3 |
| 049 | Phase II | Neurology | 4 | 4 | 4 | 3 | 7 | 10 | 3 | 4 | 3 | 4 | 2 | 2 | 6 | 6 | 4 |
| 050 | Phase II | General Surgery | 4 | 4 | 3 | 4 | 9 | 10 | 4 | 4 | 4 | 4 | 1 | 4 | 8 | 8 | 4 |
| 051 | Phase II | General Surgery | 4 | 4 | - | - | 9 | 9 | - | 3 | - | 3 | - | 4 | - | 7 | 4 |
| 052 | Phase II | Neurology | 4 | 2 | 1 | 1 | 9 | 3 | 3 | 4 | 4 | 4 | 3 | 3 | 7 | 9 | 4 |
| 053 | Phase II | Internal Medicine | 4 | 3 | 3 | 4 | 1 | 1 | 3 | 3 | 3 | 3 | 2 | 2 | 7 | 8 | 3 |
| 054 | Phase II | Neurology | 4 | 3 | 4 | 3 | 6 | 5 | 3 | 3 | 3 | 3 | 2 | 3 | 5 | 8 | 3 |
| 055 | Phase II | IM-Gastroenterology | 3 | 4 | 4 | 4 | 4 | 5 | 3 | 4 | 3 | 3 | 3 | 4 | 7 | 8 | 4 |
| 056 | Phase II | Neurology | 4 | 3 | 3 | 3 | 7 | 5 | 3 | 4 | 3 | 3 | 3 | 3 | 7 | 8 | 4 |
| 057 | Phase II | Orthopedics | 4 | 3 | - | - | 9 | 10 | 2 | 4 | 2 | 4 | 2 | 4 | 5 | 8 | 4 |
| 058 | Phase II | Internal Medicine | 4 | 4 | 4 | 4 | 8 | 8 | 4 | - | 3 | - | 2 | - | 7 | - | 3 |
| 059 | Phase II | Internal Medicine | 4 | 4 | 2 | 2 | 8 | 10 | 2 | - | 2 | - | 2 | - | 7 | - | 3 |
| 060 | Phase II | General Surgery | 3 | 3 | 3 | 3 | 8 | 7 | 3 | - | 3 | - | 2 | - | 5 | - | 3 |
| 061 | Phase II | Neurology | 3 | 4 | - | - | 8 | 8 | 1 | - | 3 | - | 3 | - | 7 | - | 3 |
| 062 | Phase II | Orthopedics | 4 | 4 | 1 | 1 | 10 | 10 | 3 | - | 3 | - | 2 | - | 6 | - | 4 |
| 063 | Phase II | IM-Oncology | 4 | 4 | 4 | 4 | 8 | 8 | 3 | - | 3 | - | 1 | - | 7 | - | 1 |
| 064 | Phase II | Neurology | 4 | 3 | 2 | 1 | 2 | 7 | 3 | - | 3 | - | 2 | - | 5 | - | 4 |
| 065 | Phase II | Neurology | 4 | 4 | 4 | 4 | 8 | 8 | 3 | - | 2 | - | 1 | - | 5 | - | 3 |
| 066 | Phase II | Orthopedics | 3 | 3 | 3 | 3 | 7 | 8 | 4 | - | 4 | - | 3 | - | 8 | - | 4 |
| 067 | Phase II | IM-Cardiology | 4 | 4 | 1 | 1 | 7 | 7 | 3 | - | 2 | - | 2 | - | 6 | - | 4 |
| 068 | Phase II | Internal Medicine | 3 | 3 | 3 | 3 | 6 | 7 | 2 | - | 3 | - | 3 | - | 6 | - | 4 |
| 069 | Phase II | Internal Medicine | 4 | 4 | 4 | 4 | 10 | 10 | 3 | - | 3 | - | 3 | - | 5 | - | 4 |
| 070 | Phase II | Internal Medicine | 4 | 4 | 2 | 3 | 7 | 7 | 1 | - | 1 | - | 1 | - | 10 | - | 4 |
| 071 | Phase II | General Surgery | 4 | 3 | 2 | 3 | 9 | 8 | 4 | - | 4 | - | 4 | - | 3 | - | 4 |
| 072 | Phase II | Internal Medicine | 3 | 4 | 4 | 3 | 8 | 7 | 3 | - | 3 | - | 3 | - | 8 | - | 3 |
| 073 | Phase II | Neurology | 4 | 4 | 4 | 4 | 9 | 9 | 2 | - | 2 | - | 2 | - | 3 | - | 2 |
| 074 | Phase II | Neurology | 4 | 4 | 4 | 4 | 9 | 9 | 3 | - | 3 | - | 2 | - | 6 | - | 3 |
| 075 | Phase II | IM-Cardiology | 2 | 4 | 2 | 3 | 3 | 3 | 3 | - | 3 | - | 2 | - | 5 | - | 4 |
| 076 | Phase II | Neurology | 4 | 3 | 4 | 4 | 2 | 9 | 2 | - | 2 | - | 1 | - | 5 | - | 4 |
| 077 | Phase II | ENT | 3 | 3 | 3 | 4 | 7 | 7 | 3 | - | 2 | - | 1 | - | 5 | - | 4 |
| 078 | Phase II | IM-Gastroenterology | 4 | 4 | 4 | 4 | 10 | 10 | 3 | - | 3 | - | 3 | - | 7 | - | 3 |
| 079 | Phase II | ENT | 4 | 4 | 3 | 2 | 10 | 10 | 3 | - | 1 | - | 1 | - | 5 | - | 4 |
| 080 | Phase II | Neurology | 4 | 4 | 2 | 3 | 7 | 7 | 3 | - | 2 | - | 2 | - | 6 | - | 4 |
| 081 | Phase II | Neurology | 3 | 3 | 3 | 3 | 8 | 8 | 4 | - | 3 | - | 4 | - | - | - | 3 |

**Supplemental Figure 1.** ED-physician average Likert score distribution

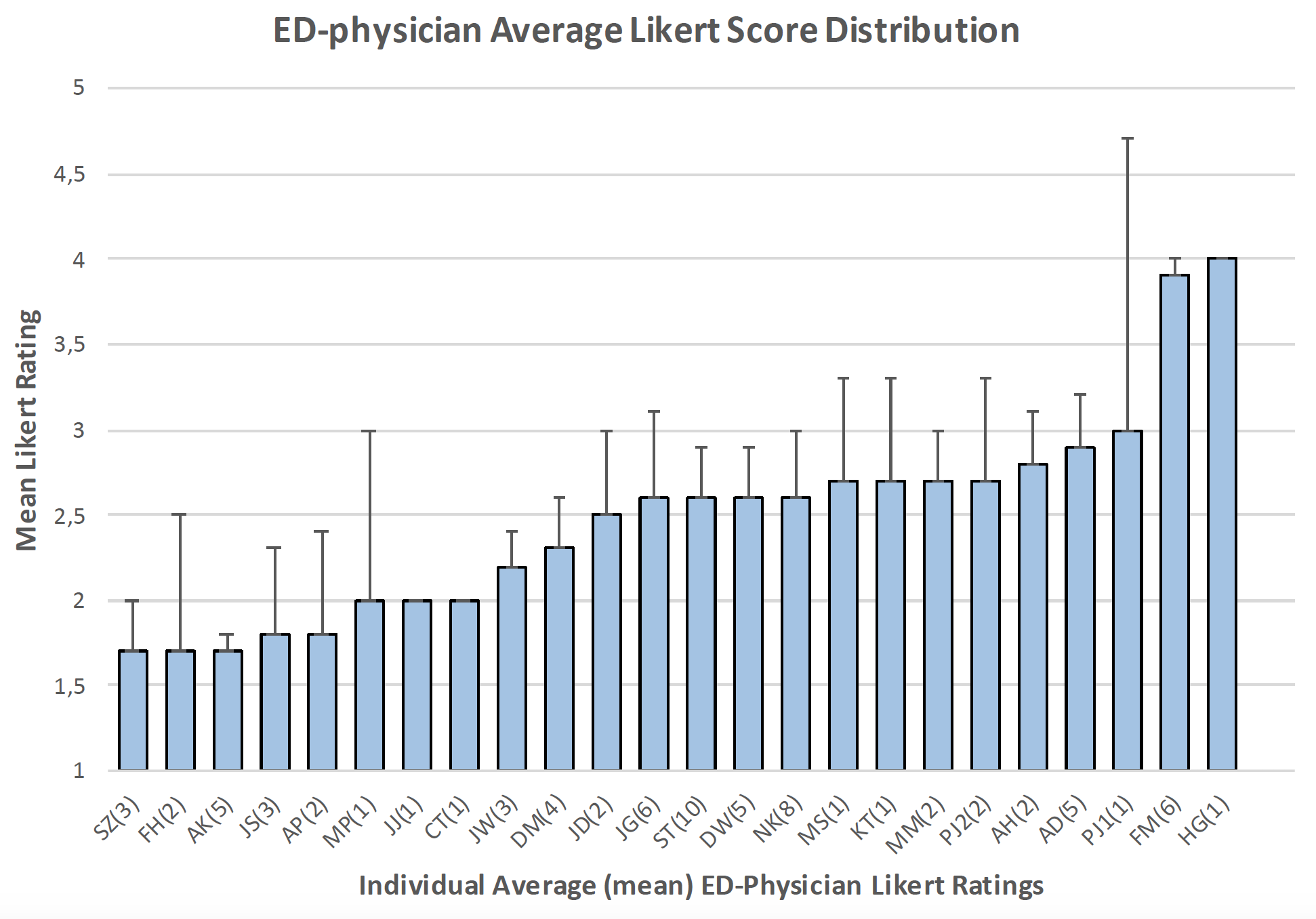

**Supplementary Table 1** This study was conducted in accordance with the requirements of the SQUIRE ^11^ and eCONSORT ^12^ guidelines, as described in this checklist).

|  | **SQUIRE** ^11^ | **Checklist and notes.** | **eCONSORT** ^12^ | **Checklist and notes.** |
| --- | --- | --- | --- | --- |
| **Title and Abstract** | **Title**  Indicate that the manuscript concerns an initiative to improve healthcare (broadly defined to include the quality, safety, effectiveness, patient-centeredness, timeliness, cost, efficiency, and equity of healthcare). | ✓ | Indicate that the intervention involves artificial intelligence/machine learning in the title and/or abstract and specify the type of model. | ✓ - AI included in the title and type of method (Bayesian reasoning engine) specified in the methods. |
|  | **Abstract**  a. Provide adequate information to aid in searching and indexing.  b. Summarize all key information from various sections of the text using the abstract format of the intended publication or a structured summary such as: background, local problem, methods, interventions, results, conclusions. | ✓ | State the intended use of the AI intervention within the trial in the title and/or abstract. | ✓ - AI included in the title and type of method (probabilistic reasoning engine) specified in the methods. |
| **Introduction** | **Problem Description**  Nature and significance of the local problem | ✓ | Explain the intended use of the AI intervention in the context of the clinical pathway, including its purpose and its intended users (e.g. healthcare professionals, patients, public). | ✓ |
|  | **Available knowledge** Summary of what is currently known about the problem, including relevant previous studies. | ✓ |  |  |
|  | **Rationale**  Informal or formal frameworks, models, concepts, and/or theories used to explain the problem, any reasons or assumptions that were used to develop the intervention(s), and reasons why the intervention(s) was expected to work. | ✓ |  |  |
|  | **Specific Aims**  Purpose of the project and of this report. | ✓ |  |  |
| **Methods** | **Context**  Contextual elements considered important at the outset of introducing the intervention(s). | ✓ | - | - |
|  | - | **-** | **Participants:** State the inclusion and exclusion criteria at the level of participants. | ✓ |
|  | - | **-** | **Participants:** State the inclusion and exclusion criteria at the level of the input data. | ✓ |
|  | - | **-** | **Participants:** Describe how the AI intervention was integrated into the trial setting, including any onsite or offsite requirements. | ✓ |
|  | **Intervention(s):** Description of the intervention(s) in sufficient detail that others could reproduce it | ✓ | **Intervention(s):** State which version of the (ii) AI algorithm was used. | ✓ |
|  | **Intervention(s):** Specifics of the team involved in the work | ✓ | **Intervention(s):** Describe how the input data were acquired and selected for the AI intervention. | ✓ |
|  | **Study of the Intervention(s):** Approach chosen for assessing the impact of the intervention(s) | ✓ | **Intervention(s):** Describe how poor quality or unavailable input data were assessed and handled. | ✓ |
|  | **Study of the Intervention(s):** Approach used to establish whether the observed outcomes were due to the intervention(s) | ✓ | **Intervention(s):** Specify whether there was human-AI interaction in the handling of the input data, and what level of expertise was required of users. | ✓ |
|  | **-** | - | **Intervention(s):** Specify the output of the AI intervention. | ✓ |
|  | **-** | - | **Intervention(s):** Explain how the AI intervention’s outputs contributed to decision-making or other elements of clinical practice. | ✓ |
|  | **Measures**  a. Measures chosen for studying processes and outcomes of the intervention(s), including rationale for choosing them, their operational definitions, and their validity and reliability.  b. Description of the approach to the ongoing assessment of contextual elements that contributed to the success, failure, efficiency, and cost.  c. Methods employed for assessing completeness and accuracy of data. | ✓ | **-** | - |
|  | **Analysis**  a. Qualitative and quantitative methods used to draw inferences from the data.  b. Methods for understanding variation within the data, including the effects of time as a variable. | ✓ | **-** | - |
|  | **Ethical Considerations**  Ethical aspects of implementing and studying the intervention(s) and how they were addressed, including, but not limited to, formal ethics review and potential conflict(s) of interest. | ✓ | **Harms**  Describe results of any analysis of performance errors and how errors were identified, where applicable. If no such analysis was planned or done, justify why not. | ✓ |
| **Results** | Initial steps of the intervention(s) and their evolution over time (e.g., time-line diagram, flow chart, or table), including modifications made to the intervention during the project. | ✓ | **-** | **-** |
|  | Details of the process measures and outcome. | ✓ | **-** | **-** |
|  | Contextual elements that interacted with the intervention(s). | ✓ | **-** | **-** |
|  | Observed associations between outcomes, interventions, and relevant contextual elements. | ✓ | **-** | **-** |
|  | Unintended consequences such as unexpected benefits, problems, failures, or costs associated with the intervention(s). | ✓ | **-** | **-** |
|  | Details about missing data. | ✓ | **-** | **-** |
| **Discussion** | **Summary:** Key findings, including relevance to the rationale and specific aims | ✓ | **-** | **-** |
|  | **Summary:** Particular strengths of the project | ✓ | **-** | **-** |
|  | **Interpretation:** Nature of the association between the intervention(s) and the outcomes. | ✓ | **-** | **-** |
|  | **Interpretation:** Comparison of results with findings from other publications. | ✓ | **-** | **-** |
|  | **Interpretation:** Impact of the project on people and systems. | ✓ | **-** | **-** |
|  | **Interpretation:** Reasons for any differences between observed and anticipated outcomes, including the influence of context. | ✓ | **-** | **-** |
|  | **Interpretation:** Costs and strategic trade-offs, including opportunity costs. | This has not been described as this is a study of a prototype and further prototype development into an on the market product would be required before costs and trade-offs could be determined. | **-** | **-** |
|  | **Limitations:** Limits to the generalizability of the work. | ✓ | **-** | **-** |
|  | **Limitations:** Factors that might have limited internal validity such as confounding, bias, or imprecision in the design, methods, measurement, or analysis. | ✓ | **-** | **-** |
|  | **Limitations:** Efforts made to minimize and adjust for limitations. | ✓ | **-** | **-** |
|  | **Conclusions:** Usefulness of the work.  . | ✓ | **-** | **-** |
|  | **Conclusions:** Sustainability. | ✓ | **-** | **-** |
|  | **Conclusions:** Potential for spread to other contexts. | ✓ | **-** | **-** |
|  | **Conclusions:** Implications for practice and for further study in the field. | ✓ | **-** | **-** |
|  | **Conclusions:** Suggested next steps | ✓ | **-** | **-** |
| **Other information** | **-** | **-** | **Registration**  Registration number and name of trial registry. | The trial was an observational trial of a software prototype. Patients were not shown output data of the prototype and HCPs assessed the tool but did not use it to inform their history taking in the study. Study registration was not a legal requirement and is not a journal requirement for this study type. |
|  | **-** | **-** | **Protocol**  Where the full trial protocol can be accessed, if available. | The protocol is only available as described in this manuscript. |
|  | **-** | **-** | **Funding**  Sources of funding and other support (such as supply of drugs), role of funders.  State whether and how the AI intervention and/or its code can be accessed, including any restrictions to access or re-use. | ✓ |
